# Suicidality and Drug Use Behavior Among Perinatal Individuals in Recovery

**DOI:** 10.64898/2026.03.03.26347368

**Authors:** Anna Constantino-Pettit, Xiao Li, Hannah Szlyk, Erin Kasson, Patricia Cavazos-Rehg

## Abstract

**Introduction:** Maternal mental health conditions, comprising maternal suicide and drug overdose, are currently the leading cause of maternal mortality in the United States. However, the relationship between suicidality and drug use behavior in the perinatal period is not well understood. We examined the association between suicidality and drug use behavior among perinatal individuals. Given the racial disparities in both drug use and suicide rates in the U.S., we also examined any differences in suicidality and drug use behavior by race.

**Methods:** Participants were recruited from a High-Risk Obstetric & Gynecological Clinic in the Midwestern U.S that specializes in providing obstetric care to perinatal individuals who have histories or current use of opioids and other illicit drugs. Participants (N = 66) were a sub-sample of a larger cohort enrolled in an mHealth intervention to support recovery from opioid and stimulant use disorders. We performed chi-square tests and t-tests to examine any significant associations between lifetime suicidality and drug use behavior during the perinatal period.

**Results:** The final analytic sample included participants who had responded to the suicidality survey questions (n=43). Nearly 40% (n=16) of our sample endorsed a lifetime history of suicidal thoughts and behaviors (SITB). Of those, 87% (n=15) reported a previous suicide *attempt*. SITB was significantly associated with cravings for opioids during the perinatal period (p = .01) as well as comorbidities with perinatal anxiety symptoms? ( p < .05), depression symptoms? (p < .05), and bipolar disorder (p < .05). A higher proportion of recent cannabis use was found among mothers with SITB, compared to those without SITB (p=0.04). Mothers with SITB also had a strong positive correlation between preconception and postnatal nicotine use compared to mothers without SITB (p < .01). Finally, while white mothers endorsed more lifetime overdoses (p= 0.01), Black mothers endorsed higher cravings for opioids during pregnancy (p = 0.03).

**Conclusions:** A history of SITB is a distinct risk factor for both illicit and recreational drug use behavior in the perinatal period, and frequently co-occurs with other perinatal mental health conditions. Further research is needed to better understand the directionality of this relationship and the complex interplay between high risk drug use behavior and suicidality.

## Introduction

The opioid epidemic has contributed to a rise in opioid use disorders (OUDs) among childbearing and postpartum individuals, with rates of OUDs among this population increasing by a staggering 131% from 2010 to 2017 (Kroelinger et al., 2019). The vast majority of research on OUDs during pregnancy has focused on the impact to the developing fetus or newborn.

Opioid use in pregnancy has most commonly been associated with adverse neonatal outcomes such as neonatal opioid withdrawal syndrome (NOWS), low birth weight, and neonatal hospitalization (Piske et al., 2021; Tobon et al., 2019). Relatively less attention has been paid to the effects of perinatal OUDs on the childbearing individual themselves, particularly from a psychiatric standpoint. A recent review by Arnaudo et al. (2017) concluded that longitudinal, population-based studies dedicated to psychiatric and substance use comorbidity during the perinatal period were critical to understanding the true prevalence of psychiatric comorbidities among perinatal individuals with OUDs. The existing studies that have examined psychiatric burden among perinatal individuals with OUDs found anxiety and depression to be the most common comorbid disorders (Shen et al. 2020; Rogers et al., 2021).

A significant protective factor against adverse behavioral health, substance-related, and neonatal outcomes is using medications for opioid use disorder, or MOUD, during the perinatal period. However, there are significant racial disparities in the use of MOUD during pregnancy, with Black non-Hispanic and Hispanic women utilizing MOUD at significantly lower rates than their white counterparts in one population-based study (Schiff et al., 2020; Austin et al., 2023). Inequitable access to MOUD in turn predisposes individuals to higher risk psychiatric and substance-related outcomes. To address these barriers, many obstetric clinics have adopted co-located models of care to provide perinatal individuals with MOUD, and have demonstrated favorable results including decreased hospital admissions and maintained recovery for childbearing individuals (Krans et al., 2021). However, the rise of these types of programs has coincided with a wave of legislation mandating reporting of childbearing individuals using MOUD to the child welfare system. This places minoritized childbearing individuals in a uniquely vulnerable situation given the history of surveillance within the child welfare system towards communities of color (Roberts, 2022; Work et al., 2023). Consequently, there continue to be structural barriers to evidence-based treatment for opioid use disorder during pregnancy (such as), which negatively impacts both recovery potential and mental health comorbidities.

A separate but related phenomenon affecting perinatal individuals is the growing burden of perinatal suicide (De Backer, 2023). Despite evidence that the prevalence of perinatal suicide has increased in recent years, inconsistencies – or even complete lack of – reporting on perinatal suicide has made it extremely difficult to understand the true burden of suicidality among this population. Risk factors for perinatal suicide include family history of suicide, prior suicidal ideation, and a prior suicide attempt (Reid et al., 2022). There are also racial and ethnic differences in rates and types of suicidality; while Black, Indigenous, and People of Color (BIPOC) women are more greatly impacted by suicidal ideation, non-Hispanic white women have a higher risk of suicide (Tabb et al. 2020). It appears that suicide risk often increases during the late postpartum period, with one prior study reporting increases in maternal suicides between six and twelve months postpartum and another documenting increases following the ‘fourth trimester’ (three months after giving birth; Petersen et al., 2019; Goldman-Mellor et al., 2019). Relatedly, studies of perinatal individuals with OUDs have reported increases in drug use during the postpartum period (Schiff et al., 2021), making the postpartum time a highly vulnerable phase for individuals experiencing both suicidality and urges for drug use.

Historically, both the research literature and clinical approaches to perinatal suicidality versus perinatal drug use have shared only a modest overlap. This has changed with the latest data showing that *mental health conditions* - comprising drug overdose and suicide - are now the leading cause of maternal mortality in the U.S., and a public health emergency (Trost et al., 2021). The rate of drug-related perinatal deaths alone increased 190% from 2010 – 2019 (Margerison et al., 2022). In the general population, opioid prescriptions per capita have also been significantly correlated with suicide deaths in recent years (Olfson et al., 2023), indicating that the comorbid burden of suicidality and opioid use disorders extends beyond perinatal individuals. Current estimates of comorbid SITBs and OUD in the general population hover around 30%, although this is believed to be an underestimate (Na et al, 2022).

Disentangling the complex relationship between suicidality and drug overdose in the perinatal population remains a challenge. While 49 out of 50 states in the U.S. have established maternal mortality review committees to review pregnancy-related deaths, only 36 states review deaths that happen during the first postpartum year – despite the fact that maternal mortality typically encompasses death by a childbearing or postpartum individual up to one year postpartum (Centers for Disease Control and Prevention, 2023; World Health Organization, 2023). Additionally, the suicide rate in the U.S. is highest among adults ages 25-34 – during female-identified individuals’ prime reproductive years (American Foundation for Suicide Prevention, 2023). While the burden of suicide is high among perinatal individuals, the association between suicidality and drug use behaviors during the perinatal period is not well understood.

The current study had two aims. First, it attempts to characterize the burden of lifetime suicidality among perinatal individuals with OUDs. Second, it additionally examines any differences by race in mental health comorbidities and substance use behaviors during the perinatal period. Understanding the relative contributions of suicidality and race to perinatal drug use and mental health comorbidities can inform the development of equitable interventions that address both substance use and mental health needs, ultimately promoting better outcomes for both the childbearing individual and their child.

## Materials and Methods

### Participants

Participants for the current study were recruited from a Midwestern, urban, high-risk obstetrics and gynecological (OB-GYN) clinic that specializes in providing prenatal care to childbearing individuals with either OUDs or other drug and alcohol related disorders.

Participants (n=66) enrolled in the current study had consented to participate in a mobile health (“mHealth”) intervention for one month (see Cavazos-Rehg et al., 2020, for a full description of the intervention). Eligibility for enrollment included: having a diagnosis of an OUD, being either currently pregnant or within one year postpartum, being at least 18 or older, being a U.S. resident, English language fluency, and owning a smartphone with an Apple or Android operating system.

### Procedure

From 2018 to 2021, trained research staff screened individuals for eligibility via the electronic medical record (EMR) and contacted prospective participants via phone to review the informed consent and study objectives. Participants provided either verbal or written consent to be enrolled in the study. Participants also gave consent to access some information directly from the EMR; research staff collected participant demographic information, current diagnostic codes, urine drug screens, gravidity, and parity. All data was stored in Research Electronic Data Capture (REDCap). Participants were then administered REDCap surveys around the time of enrollment (“Baseline”) and at their 1-month follow-up through the *u-MAT-R* app. For the purposes of the current study, only participants who had completed the section of the baseline survey pertaining to suicidality in its entirety (n=43) were included. This study was approved by the Washington University Institutional Review Board (IRB #201805132).

### Study variables

**Demographic covariates.** All participants were asked to self-identify their gender (options: Male, Female, Transgender, Non-binary, Genderqueer, Other). Age was calculated from the participants’ birthdates which were harvested from the EMR. Race/ethnicity data was also harvested from the EMR; options included “Yes, Hispanic or Latino, No, not Hispanic or Latino, White, Black or African-American, American-Indian or Alaskan Native, Asian, Native Hawaiian or Pacific Islander, and From multiple races”. Insurance status was harvested from the medical record; participants were asked whether they were on public health insurance, private health insurance, or other. Employment status was ascertained via participant self-report (“Are you currently employed?”) with the following response options: “Yes, full-time”; “Yes, part-time”; “No, I am not employed”. These responses were recoded into a binary variable with 1 = full or part time employment and 0 = unemployed. Finally, pregnant individuals’ gestational age and estimated delivery date (EDD) were recorded from the EMR. If postpartum, the number of days postpartum was calculated using the delivery date from the EMR. This data was collapsed and recoded into either ‘Pregnant’ or ‘Postpartum’.

### Mental health comorbidities

*Medical record data*. Active mental health diagnostic codes were lifted from the EMR at the time of the baseline survey distribution. These diagnoses were imported from REDCap and recoded into the following categories: Bipolar Disorder (‘0’ = No; ‘1’ = Yes) and Posttraumatic Stress Disorder (‘0’ = No; ‘1’ = Yes). Some participants also had diagnoses of anxiety and depression reflected in their EMR. However, because there are frequently structural barriers associated with receiving a formal diagnosis for anxiety and depression (e.g., lack of access to mental healthcare; insufficient screening in primary care settings), we relied on participant self-report in an effort to capture a more accurate depiction of perinatal anxiety and depression.

*Self-report data.* Participants completed the Edinburgh Postnatal Depression Scale (EPDS; Cox et al., 1987) at the time of the baseline survey. EPDS scores were calculated and used as a continuous variable (“raw score”). Participants additionally completed the 7-item Generalized Anxiety Scale (GAD-7; Spitzer et al., 2006). Results from the GAD-7 were similarly scored and used as a continuous variable.

**Suicidality.** Participants answered the following questions pertaining to suicidal history: “Has there been a time in the past month when you have had serious thoughts about ending your life?” (1= Yes; 0 = No), and “Have you ever, in your whole life, tried to kill yourself or made a suicide attempt?” (1 = Yes; 0 = No). This data was aggregated into a single variable for suicidal thoughts and behaviors (SITB; 1 = Yes; 0 = No).

**Opioid Use.** Participants answered a series of questions about opioid use, both over the course of the past month and current cravings at the time of the baseline survey. Participants reported lifetime overdose count (“How many times have you overdosed on drugs?”). Participants additionally answered a series of questions about cravings for opioids: “How much do you currently crave opioids?” (in Table 3: ‘*Current cravings for opioids – baseline’*); “In the past week, please rate how strong your desire to use opioids has been when something in the environment has reminded you of opioids” (in Table 3: ‘*Cue cravings’*); and “Please imagine yourself in the environment in which you previously used opioids. If you were in this environment today and if it were the time of day that you typically used opioids, what is the likelihood that you would use opioids today?” (in Table 3: ‘*Likelihood of opioid use – given means’*). For each question, participants rated their craving or likelihood from 0 (Not at all) to 10 (extremely). These items were retained and used as a continuous variable for all analyses related to opioid cravings.

**Other drug and alcohol use.** Participants were also queried on both lifetime (preconception) and perinatal (past-month) nicotine, alcohol, and cannabis use. Perinatal substance use was stored as an ordinal-level variable with the following values: 0 – *Not at all*; 1 – *A few times*; 2 – *A few times each week*; 3 – *Every day*. Preconception substance use was stored as an ordinal-level variable with the following values: 0 – *0 occasions*; 1 – *1-2 occasions*; 2 – *3-5 occasions*; 3 – *6-9 occasions*; 4 – *10-19 occasions*; 5 – *20-39 occasions*; 6 – *40 or more occasions*. For both the perinatal and preconception substance use variables, responses were retained as recorded and analyzed as continuous variables. Finally, a polysubstance use variable was derived by summing the responses to the past-month nicotine, alcohol, and cannabis use questions for each participant.

### Missing variables

Due to the modest sample size, a complete case analysis was favored over imputation for the handling of missing data. The analytic sample size, 43, was comprised of individuals who had completed the entire baseline survey including each of the questions pertaining to suicidality. There were no significant differences on key demographic factors (age, race, and education) between those who had completed the baseline survey and those who had not. See Table 1 in the Supplement for results of the t-tests and chi-square tests.

**Table 1.** Sociodemographic characteristics by lifetime history of suicidal thoughts and behaviors (SITB)

|  | All<br>(n=43) | No SITB<br>(n=27)<br>M (SD) or N (%) | SITB<br>(n=16) | X<sup>2</sup>/t-value | p |
| --- | --- | --- | --- | --- | --- |
| Age | 29.2 (4.4) | 29.7 (3.9) | 29.1 (4.9) | 0.42 | 0.68 |
| Race |  |  |  |  |  |
| White | 29 (67.4) | 19 (70.3) | 10 (62.5) | 0.04 | 0.84 |
| Black | 14 (32.6) | 8 (29.6) | 6 (37.5) |  |  |
| Perinatal Status |  |  |  |  |  |
| Pregnant | 34 (79.0) | 21 (77.8) | 13 (81.2) | -- | 1 |
| Postpartum | 9 (20.9) | 6 (22.2) | 3 (18.8) |  |  |
| Marital Status |  |  |  |  |  |
| Married | 8 (18.6) | 6 (22.2) | 2 (12.5) | -- | 0.69 |
| Single | 35 (81.4) | 21 (77.8) | 14 (87.5) |  |  |
| Employment |  |  |  |  |  |
| Full/Part time | 9 (20.9) | 6 (22.2) | 3 (18.8) | -- | 1 |
| Not employed | 34 (79.0) | 21 (77.8) | 13 (81.2) |  |  |
| Insurance status |  |  |  |  |  |
| Medicaid | 41 (95.3) | 27 | 14 | -- | 0.13 |
| Private | 2 (0.05) | 0 | 2 |  |  |

### Statistical analyses

Descriptive statistics were performed on the complete analytic sample (n=43). T-test (continuous) and chi-square (categorical) tests were run to examine differences by lifetime suicidality status on the following demographic characteristics: perinatal status, marital status, race, age, insurance type, and employment. T-tests then examined differences between those with and without a lifetime history of suicidality on the following characteristics: past month cravings for opioids; current cravings for opioids; likelihood of use; EPDS score at baseline; GAD score at baseline; lifetime overdose count; polysubstance use; and preconception and perinatal nicotine, alcohol, and cannabis use. A chi-square test (or fisher’s exact test, in the case of low cell counts; Fisher, 1922) was used to analyze differences in rates of bipolar disorder and posttraumatic stress disorder between those with and without a history of SITB. A subsequent analysis examined differences in mental health and drug use characteristics between blackand white participants. Any mental health or drug use characteristics that were found to differ significantly between white and black participants were then entered into a separate analysis of covariance (ANCOVA) test to determine the relative contribution of race versus SITB on the outcome of interest. Finally, we performed correlation tests on rates of preconception versus perinatal nicotine, cannabis, and alcohol use, stratifying by SITB status. All analyses were conducted using R and RStudio (R Core Team 2023).

## Results

### Suicidal ideation, thoughts, and behaviors

Nearly 40% (n=16) of participants endorsed SITBs. Of those, a majority reported a prior suicide attempt in their lifetime (n=15; 94%), while 2 (13%) reported suicidal ideation in the past month. Suicide attempt was queried distinct from lifetime history of overdose so that accidental overdose was not misconstrued as overdose with intent to die. Participants were not queried about means involved in their suicide attempt; while some attempts may have involved drug misuse, it is also possible that there were other means utilized.

### Demographic characteristics

Demographic characteristics of the analytic sample are presented in Table 1. Participants were, on average, 29 years old, and all participants identified as female. The majority of the sample was single (n= 35; 81%), white (n=29; 70%), and unemployed (n=34; 79%). Most participants (n= 34; 79%) were pregnant at the time of the baseline survey, although a handful of women (n=9; 20%) were postpartum (average postpartum days = 148). While all women were insured, the vast majority (n=41; 95%) were on public insurance. There were no significant differences by SITB on any of the demographic characteristics in this sample.

### Suicidality, opioid use, and other mental health comorbidities

Table 2 presents the results of differences in mental health comorbidities and drug use behavior by SITB. Perinatal depression symptoms (t(33)= −2.2, p= 0.04), perinatal anxiety symptoms (t(18)= −2.7, p= 0.02), and bipolar disorder (Fisher’s exact test; p= .04) were all significantly higher among the participants with SITB. Posttraumatic stress disorder was the only mental health comorbidity to not differ significantly based on SITB. All three self-reported cravings scores were significantly higher among those with SITB. Participants with SITB rated their current cravings (t(27)= −2.5, p= 0.02), past week cravings (t(24)= −2.4, p= 0.03), and likelihood of use (t(22)= −2.6, p= 0.02) as significantly higher than those without SITB. There were no significant differences in drug overdoses by SITB in this sample.

**Table 2.**
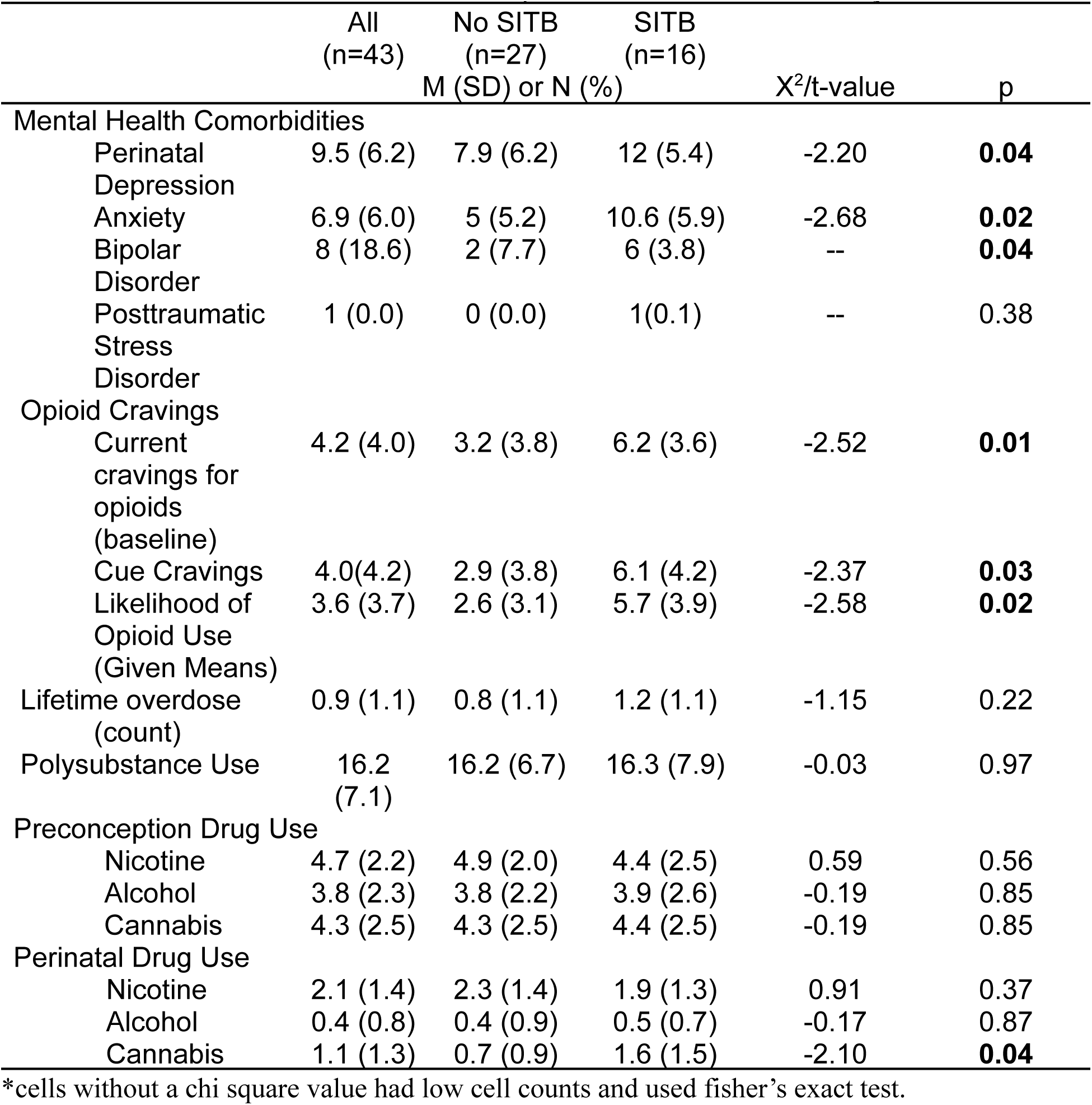
Mental health comorbidities and past month substance use by lifetime SITB.

|  | All<br>(n=43) | No SITB<br>(n=27) | SITB<br>(n=16) |  |  |
| --- | --- | --- | --- | --- | --- |
|  | M (SD) or N (%) |  |  | X<sup>2</sup>/t-value | p |
| Mental Health Comorbidities |  |  |  |  |  |
| Perinatal Depression | 9.5 (6.2) | 7.9 (6.2) | 12 (5.4) | -2.20 | **0.04** |
| Anxiety | 6.9 (6.0) | 5 (5.2) | 10.6 (5.9) | -2.68 | **0.02** |
| Bipolar Disorder | 8 (18.6) | 2 (7.7) | 6 (3.8) | -- | **0.04** |
| Posttraumatic Stress Disorder | 1 (0.0) | 0 (0.0) | 1(0.1) | -- | 0.38 |
| Opioid Cravings |  |  |  |  |  |
| Current cravings for opioids (baseline) | 4.2 (4.0) | 3.2 (3.8) | 6.2 (3.6) | -2.52 | **0.01** |
| Cue Cravings | 4.0(4.2) | 2.9 (3.8) | 6.1 (4.2) | -2.37 | **0.03** |
| Likelihood of Opioid Use (Given Means) | 3.6 (3.7) | 2.6 (3.1) | 5.7 (3.9) | -2.58 | **0.02** |
| Lifetime overdose (count) | 0.9 (1.1) | 0.8 (1.1) | 1.2 (1.1) | -1.15 | 0.22 |
| Polysubstance Use | 16.2 (7.1) | 16.2 (6.7) | 16.3 (7.9) | -0.03 | 0.97 |
| Preconception Drug Use |  |  |  |  |  |
| Nicotine | 4.7 (2.2) | 4.9 (2.0) | 4.4 (2.5) | 0.59 | 0.56 |
| Alcohol | 3.8 (2.3) | 3.8 (2.2) | 3.9 (2.6) | -0.19 | 0.85 |
| Cannabis | 4.3 (2.5) | 4.3 (2.5) | 4.4 (2.5) | -0.19 | 0.85 |
| Perinatal Drug Use |  |  |  |  |  |
| Nicotine | 2.1 (1.4) | 2.3 (1.4) | 1.9 (1.3) | 0.91 | 0.37 |
| Alcohol | 0.4 (0.8) | 0.4 (0.9) | 0.5 (0.7) | -0.17 | 0.87 |
| Cannabis | 1.1 (1.3) | 0.7 (0.9) | 1.6 (1.5) | -2.10 | **0.04** |

\*cells without a chi square value had low cell counts and used fisher's exact test.

**Table 3.** Mental health comorbidities and drug use behavior by Race.

|  | All<br>(n=43) | White<br>(n=29) | Black or<br>African<br>American<br>(n=14) |  |  |
| --- | --- | --- | --- | --- | --- |
|  | M (SD) or N (%) |  |  | X<sup>2</sup>/t-value | p |
| SITB | 16 (37.2) | 10 (34.5) | 6 (42.9) | 0.04 | 0.84 |
| Mental Health Comorbidities |  |  |  |  |  |
| Perinatal Depression | 9.5 (6.2) | 9.0 (6.5) | 10.3 (5.7) | -1.25 | 0.22 |
| Anxiety | 6.9 (6.0) | 6.8 (6.6) | 7.1 (4.9) | -0.16 | 0.88 |
| Bipolar Disorder | 8 (18.6) | 4 (14.3) | 4 (28.6) | -- | 0.41 |
| Posttraumatic Stress Disorder | 1 (0.0) | 0 (0.0) | 1(0.1) | -- | 0.38 |
| Opioid Cravings |  |  |  |  |  |
| Current cravings for opioids (baseline) | 4.2 (4.0) | 3.3 (4.0) | 6.0 (3.3) | -2.33 | **0.03** |
| Cue Cravings | 4.0(4.2) | 3.1 (4.0) | 5.6 (4.1) | -1.89 | 0.07 |
| Likelihood of Opioid Use (Given Means) | 3.6 (3.7) | 3.3 (3.7) | 4.4 (3.8) | -2.58 | 0.40 |
| Lifetime overdose (count) | 0.9 (1.1) | 1.2 (1.2) | 0.4 (0.6) | 2.66 | **0.01** |
| Polysubstance Use | 16.2 (7.1) | 16.3 (7.6) | 16.1 (6.3) | 0.10 | 0.92 |
| Preconception Drug Use |  |  |  |  |  |
| Nicotine | 4.7 (2.2) | 4.8 (2.3) | 4.6 (2.0) | 0.23 | 0.82 |
| Alcohol | 3.8 (2.3) | 4.1 (2.3) | 3.3 (2.5) | 0.94 | 0.36 |
| Cannabis | 4.3 (2.5) | 4.4 (2.5) | 4.2 (2.6) | 0.29 | 0.77 |
| Perinatal Drug Use |  |  |  |  |  |
| Nicotine | 2.1 (1.4) | 1.9 (1.5) | 2.5 (1.1) | -1.41 | 0.17 |
| Alcohol | 0.4 (0.8) | 0.5 (0.9) | 0.4 (0.6) | 0.52 | 0.61 |
| Cannabis | 1.1 (1.3) | 1.0 (1.2) | 1.1 (1.4) | -0.25 | 0.81 |

### Other drug use

Differences in preconception or perinatal alcohol, nicotine, and cannabis use by suicide history are also presented in Table 2. Perinatal cannabis use alone was significantly higher among those with a history of SITB (t(22)= −2.10, p=0.04). Polysubstance use (a composite of the past-month and lifetime nicotine, cannabis, and alcohol use) did not differ significantly by SITB. We additionally examined the correlations between rates of preconception drug use and perinatal drug use among those with versus without a history of suicidality. The relationships between preconception and perinatal drug use are depicted in Figure 1. The correlation between preconception and perinatal nicotine use was statistically significant among individuals with a history of suicide (r = 0.73, p < .01) but not for those without a history of suicide (r = 0.03, p = 0.89). A Fisher r-to-z transformation determined that the differences between the two correlations was also significant (z = −2.65, p = 0.01). Neither the correlation between preconception and perinatal alcohol use for individuals with a history of suicide (r = −0.02, p = 0.94) nor those without a history of suicide (r = 0.16, p = 0.44) was statistically significant. Similarly, the correlation between preconception and perinatal cannabis use was not significant for those both with (r = 0.24, p = 0.41) and without (r = 0.22, p = 0.29) a history of suicide.

**Figure 1.**
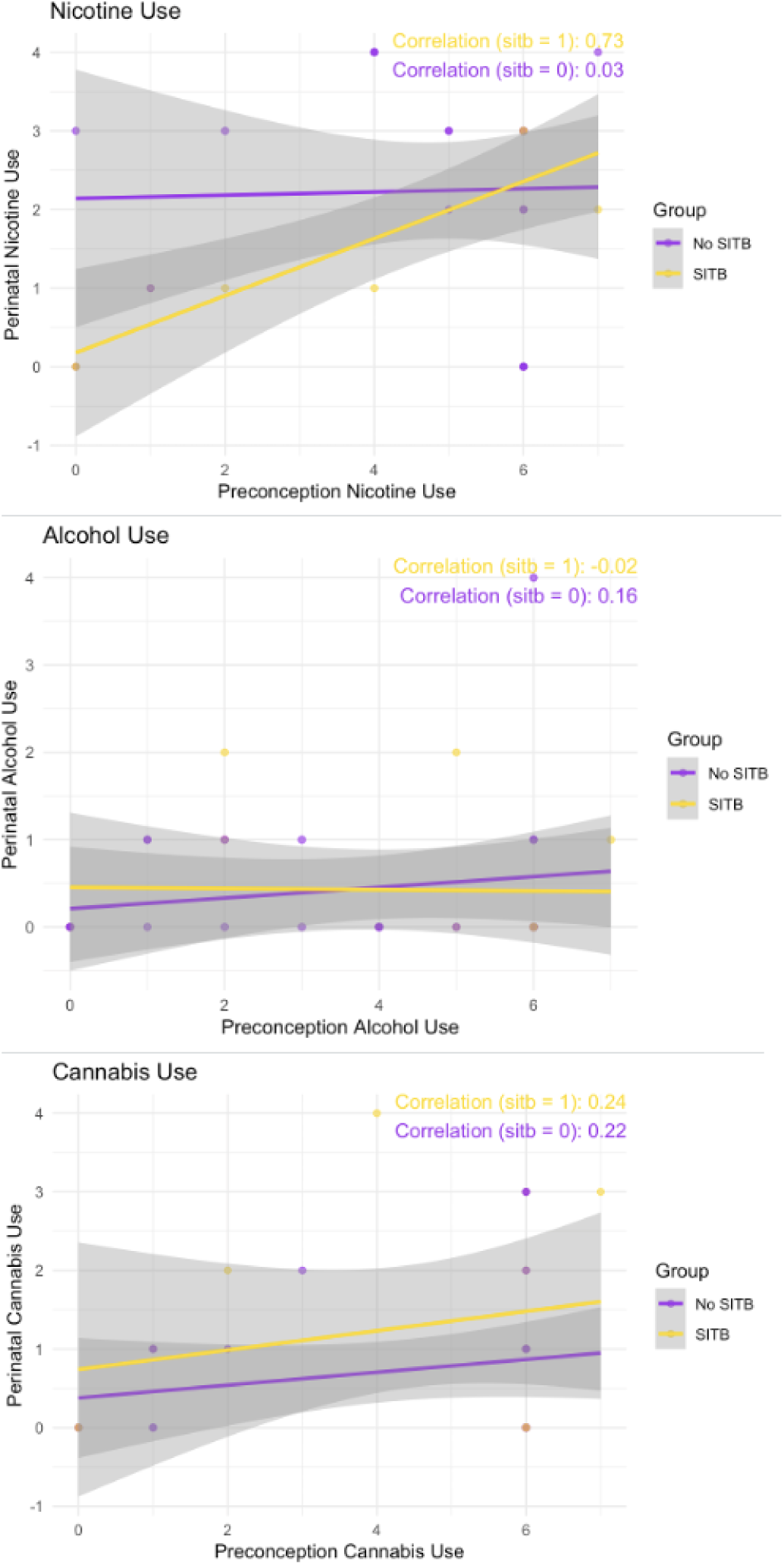
**Correlations Between Preconception and Perinatal Substance Use by Mothers With and Without SITB**

### Race-related differences in mental health and drug use behavior

Table 3 presents results of differences in mental health comorbidities and drug use behavior by race. We were interested in examining differences by race separately from the analyses by SITB in order to identify any race-related effects driving the association between SITB and the outcome variables of interest. While SITBs did not differ by race, lifetime overdose count was significantly associated with race, with white mothers reporting more lifetime overdose events compared to black mothers (t(39)= 2.66, p= 0.01) . Current cravings also differed significantly by race: in contrast to the lifetime overdose findings, black mothers reported higher current cravings compared to their white peers (t(31)= −2.33, p = 0.03). No other mental health comorbidities or drug use behaviors were significantly associated with race.

### Relative contributions of race-related factors and SITB to current cravings

Stratifying the analytic sample by race resulted in lifetime overdose count being significantly related to race but not history of SITB. In contrast, both race and history of SITB were significantly associated with current cravings. We performed an analysis of covariance (ANCOVA) to better understand the relative contribution of both SITB and race-related factors to current cravings. Race was added into the model first in order to discern any additional effect of SITB history on current cravings. The results of the ANCOVA are displayed in Table 4. While race remained a significant predictor of current cravings, SITB maintained a larger effect (*F*(1,38)=6.70, *p*=0.01, *η*^2^=0.15) and therefore appeared to be primarily driving the significant differences in rates of current cravings.

**Table 4.** Analysis of Covariance for Cravings by Suicidal History, Controlling for Race.

| Dependent Variable | Source | df | SS | MS | F | p | $\eta^2$ |
| --- | --- | --- | --- | --- | --- | --- | --- |
| Current Cravings | Race | 1 | 51.2 | 51.2 | 3.9 | **0.05** | 0.09 |
|  | SITB | 1 | 86.7 | 86.7 | 6.7 | **0.01** | 0.15 |
|  | Error | 38 | 494.6 | 13.0 | -- | -- |  |

## Discussion

The current study investigated the relationship between lifetime suicidality and drug use behavior in perinatal individuals. The results indicated that individuals with a history of suicidality had significantly higher cravings and perinatal mental health comorbidities – with the exception of posttraumatic stress disorder - compared to those without a history of suicidality.

Mothers with a history of suicidality endorsed higher rates of cannabis use during pregnancy. These mothers additionally had a strong positive correlation between preconception nicotine use and perinatal nicotine use. When examining differences by race, white mothers endorsed higher rates of lifetime overdose. In contrast, black mothers reported higher current cravings for opioids. However, additional analyses showed a history of suicide to be a greater contributor to current cravings for opioids compared with race. These findings highlight the complex relationship between suicidality, OUDs, and mental health during the perinatal period.

Our findings should be interpreted in context with other literature on psychiatric comorbidities with drug use disorders during the perinatal period. A recent review by Arnaudo et al. (2021) identified only 21 articles that described psychiatric comorbidities among pregnant women with substance use disorders. While they did not focus on suicidality, specifically, the authors concluded that pregnant women with psychiatric comorbidities were more likely to experience difficulty with treatment adherence for OUDs. One of the only case series to examine suicide and drug overdose among perinatal women was carried out using epidemiological data from Colorado between 2004 and 2012. These authors found that deaths were more concentrated in the postpartum period, and that – importantly – nearly half of the women who died had discontinued a psychotropic medication during their pregnancy (Metz et al., 2016). A similar case review utilizing the National Violent Death Reporting System from 2003 to 2007 compared perinatal women with non-pregnant women of reproductive age who had died by suicide; drug use rates were similar in both the perinatal and non-pregnant cohorts (Gold et al., 2012). A similar postmortem case review among perinatal and non-pregnant victims of suicide in the UK found that perinatal victims had higher rates of a prior alcohol misuse disorder, but did not find differences in other drug use (Khalifeh et al., 2016). While each of these studies highlights the burden of both suicide and drug use during the perinatal period, our study is among the first to examine SITBs distinct from drug use behavior during the perinatal period.

A notable finding from this analysis was the relationship between suicidality and cannabis and nicotine use. We found that mothers with a history of suicidality both increased nicotine use during pregnancy and also endorsed higher rates of cannabis use during pregnancy. This may be indicative of alternative strategies to cope with the burden of suicidal ideation or other urges during pregnancy. Prior studies that have examined risk factors for substance use during pregnancy have found that pregnant women using alcohol, tobacco, or cannabis during pregnancy were significantly more likely to also use other substances ( Passey et al., 2014).

Similarly, recent data from the Pregnancy Risk Assessment Monitoring System (PRAMS) found nicotine use to be a risk factor for cannabis use during pregnancy (Ko et al., 2020). To our knowledge, suicidality has not been investigated as a risk factor forcannabis and nicotine use during pregnancy. While there are well-documented risks to fetal health from nicotine exposure (Leonardi-Bee et al., 2008), we are still learning about the extent of the effects of cannabis exposure on fetal development (El Marroun et al., 2009). Regardless, these findings underscore the need for support to find alternative means of coping for mothers in recovery with comorbid suicidality.

Finally, we found two distinctions in drug use and behavior by race, with white mothers endorsing higher rates of lifetime overdose and black mothers endorsing higher current opioid cravings. Prior research has exposed significant inequities in accessibility to MOUD as well as differences in prescribing practices even when women of color have access to MOUD (Rosenthal et al., 2021; Gao et al., 2022). All women enrolled in this study were on MOUD as they were recruited from an obstetric clinic specializing in MOUD treatment. As such, we cannot speak to inequities in access to MOUD for this sample; however, it is important that even among women engaged in MOUD treatment, black women still reported higher levels of current cravings. It is possible that black mothers were experiencing increased stressors compared to white mothers, which may be driving higher cravings – but further qualitative research would be useful in identifying major drivers of high cravings during pregnancy for black mothers. The fact that white mothers endorsed higher rates of lifetime overdose compared to black mothers is in line with the most recent data from the CDC, which shows that white women comprise the highest proportion of pregnant women with deaths due to mental health conditions (Division of Reproductive Health, National Center for Chronic Disease Prevention and Health Promotion, 2022). However, Non-Hispanic black women and American Indian / Alaska Native women continue to comprise the largest proportion of victims to all pregnancy-related deaths (Petersen et al., 2019). Other recent research noted that pregnancy-related mortality increased with rural geographic location (Merkt et al., 2021). While this analysis did not include information on geographic location, it is possible that white mothers in our sample were also more likely to reside in more rural counties compared to the black mothers in our sample.

This study had several limitations. The modest sample size may limit the generalizability of these findings. While analyses revealed no significant differences in demographic factors between those who had completed the baseline survey in its entirety versus those who did not, a significant proportion of perinatal individuals started but did not complete the baseline survey.

Finally, because this paper utilized data from the baseline survey only, we were unable to determine any additional effects of the mHealth intervention on either suicidality or drug use behavior during the one-month intervention period.

Despite these limitations, this study contributes valuable insights into the mental health needs of perinatal individuals with OUDs. Our findings support the need for integrated interventions that concurrently address substance use, suicidality, and mental health conditions in this population. The mHealth intervention used in this study, which includes educational content, resources, and access to an “e-coach,” represents a promising strategy for providing such comprehensive support. Future research should continue to investigate how digital therapeutics can be optimized to address the complex needs of perinatal individuals with OUDs and to ultimately improve outcomes for both the individual and their child.

Overall, our study contributes to the growing body of literature urging for a more holistic approach in the treatment and management of pregnant people with SUDs. The significant lifetime histories of suicidal thoughts and behaviors, along with the amplified symptoms of depression during the perinatal period, call for an urgent reevaluation of care models. Mental health services must be an integral part of prenatal care, alongside targeted interventions for substance use, to better address the overlapping needs of this vulnerable population.

## Conclusions

This study highlights the intricate relationship between lifetime suicidality, cravings for OUDs, and mental health in perinatal individuals. The findings emphasize the importance of developing integrated interventions that address both substance use and mental health needs in this population. In order to address the needs of perinatal individuals with OUDs, we must devise interventions that are specifically designed to support those with a history of suicidal thoughts and behavior.

## Data Availability

All data produced in the present study are available upon reasonable request to the authors

---

**Supplement A.** Differences in demographic variables by completers / non-completers.

|  | All<br>(n=66) | Survey Not<br>Complete<br>(n=23) | Survey<br>Complete<br>(n=43) |  |  |
| --- | --- | --- | --- | --- | --- |
|  | M (SD) or N (%) |  |  | X<sup>2</sup>/t-value | p |
| Age | 29.2 (4.4) | 29.7 (3.9) | 29.1 (4.9) | -1.04 | 0.31 |
| Educational<br>Attainment | 13.4 (2.1) | 13.0 (2.4) | 13.8 (2.1) | 0.92 | 0.37 |
| Race |  |  |  |  |  |
| White | 43 (65.2) | 14 (60.9) | 29 (67.4) | 0.29 | 0.59 |
| Black | 23 (34.8) | 9 (39.1) | 14 (32.6) |  |  |

